# AMPD2 deficiency implicates cytosolic purine metabolism in the pathogenesis of Leigh syndrome

**DOI:** 10.1101/2025.09.17.25335877

**Authors:** Justin Simo, Alexandre Janer, Hana Antonicka, Gabrielle Macaron, André Mégarbané, Rami Massie, Erin O’Ferrall, Jason Karamchandani, Bernard Brais, Inge Meijer, Tanguy Demaret, Grant Mitchell, Valérie Triassi, Martine Tétreault, Eric Shoubridge, Roberta La Piana

**Affiliations:** The Neuro (Montreal Neurological Institute-Hospital), McGill University, Montreal, QC, Canada; Department of Neurology and Neurosurgery, McGill University, Montreal, QC, Canada; Department of Human Genetics, McGill University, Montreal, QC, Canada; Department of Neurology, Centre Hospitalier de l’Université de Montréal, Montreal, QC, Canada; Department of Neuroscience, Université de Montréal, Montreal, QC, Canada; Faculty of Medicine, Université Saint-Joseph de Beyrouth, Beirut, Lebanon; Department of Human Genetics, Gilbert and Rose-Marie Chagoury School of Medicine, Lebanese American University, Beirut, Lebanon; Institut Jérôme Lejeune, Paris, France; Division of Medical Genetics, Department of Pediatrics, Centre Hospitalier Universitaire Sainte-Justine, Université de Montréal, Montreal, QC, Canada; Department of Diagnostic Radiology, McGill University, Montreal, QC, Canada

**Author notes:** Correspondence to: Roberta La Piana, MD, PhD, Assistant Professor, Department of Neurology and Neurosurgery, The Neuro (Montreal Neurological Institute-Hospital), 3801 University Street, Montreal, QC H3A 2B4, Canada. Shared senior authorship (Eric Shoubridge, Roberta La Piana).

## Abstract

**Importance:** Cellular mechanisms underlying mitochondrial dysfunction, a hallmark feature of many neurodegenerative conditions, remain incompletely understood, and their true diversity is unknown.

**Objective:** To identify and functionally validate novel genetic variants causative of Leigh syndrome.

**Design:** We performed whole genome sequencing (WGS) and first-degree relative genotyping on two unrelated adult subjects with brain MRI abnormalities evoking Leigh syndrome. Blue native polyacrylamide gel electrophoresis (BN-PAGE) and respiratory chain enzymatic activity assays were performed to screen for respiratory complex assembly and/or oxidative phosphorylation impairments. Cells obtained from patient dermal and muscular biopsies were immortalized and later genetically corrected to evaluate cellular response to metabolic stress.

**Setting:** Subjects were recruited from The Neuro (McGill University), Rizk Hospital (Lebanese American University), and Centre Hospitalier Universitaire Sainte-Justine (University of Montreal). Research connections were established through the White Matter Rounds Network and GeneMatcher.

**Participants:** Four subjects representing three families with undiagnosed Leigh syndrome (age range 10-40 years) were ultimately recruited.

**Main outcome(s) and Measure(s):** DNA sequencing uncovered a new autosomal recessive Leigh syndrome-associated gene that was functionally validated.

**Results:** Bi-allelic pathogenic variants in *AMPD2* were detected in all subjects. BN-PAGE of patient skeletal muscle mitochondria captured an isolated complex V assembly defect in the context of heavy mTOR activation, while the accompanying enzymological assays reported decreased activities of complexes I and IV. Opposite to controls, patient-derived cell lines and muscle lacked AMPD2 protein, attributing null status to the variants detected. During metabolic challenge, only mutant cells suffered from mitochondrial hyperfusion and high-order cytosolic IMPDH2 oligomerization, implying simultaneous ATP accumulation and GTP deficiency. However, under these conditions, both complex V assembly and mTOR status in mutant cells and myotubes remained unchanged relative to the corrected lines. All mutant phenotypes observed collectively reverted upon exogenous introduction of wild-type AMPD2.

**Conclusions and Relevance:** The recognition of *AMPD2*-related Leigh syndrome (*AMPD2*-LS) as a novel entity provides strong evidence for classifying AMPD2 deficiency as a mitochondrial disease. Our data suggest that respiratory capacity is significantly modulated by AMPD2, a cytosolic enzyme selectively regulating complex V assembly through an elusive process.

**Key points:** *Question:* Do *AMPD2* mutations cause mitochondrial disease?

*Findings:* In this case series, we found that four subjects from three families with molecularly unexplained Leigh syndrome carried bi-allelic, loss-of-function variants in *AMPD2*, a gene not previously linked to mitochondrial disease. Biochemical analyses uncovered an isolated complex V assembly defect, providing diagnostic confirmation of a new entity: *AMPD2*-related Leigh syndrome (*AMPD2*-LS).

*Meaning:* The cytosolic purine cycle is a primordial determinant of oxidative phosphorylation and mitochondrial health.

## Introduction

Next-generation sequencing (NGS) has proven critical in identifying novel disease-causing genes and variants and expanding the phenotypes of rare neurogenetic disorders, especially in adults^1,2^. Among these disorders is Leigh syndrome, a rare neurodegenerative disorder caused by genetic mutations that ultimately impair mitochondrial oxidative phosphorylation (OXPHOS)^3,4^. All forms of Leigh syndrome, irrespective of onset age and symptomatology, share hallmark neuroimaging findings of progressive signal abnormalities in the striatal nuclei, thalami, brainstem, and/or cerebellum^3–5^. The classical presentation is an infantile encephalopathy characterized by developmental delay/regression, seizures, hypotonia, movement disorders, and lactic acidosis, with early childhood demise secondary to respiratory/hepatic insufficiency^3,4^. Adult-onset Leigh syndrome, in comparison, is ultra-rare and displays much greater clinical heterogeneity, imposing further diagnostic challenges^6^.

Adenosine monophosphate deaminase paralog 2 (AMPD2) is a conserved cytosolic enzyme gating the conversion of adenosine nucleotides to guanosine nucleotides. Uniquely positioned in purine biosynthesis, AMPD2 therefore balances many energy-demanding cellular processes commonly disrupted in neurodegenerative disorders^7–9^. Bi-allelic, loss-of-function variants in the *AMPD2* gene have so far been described in only 36 patients with either infantile spastic paraplegia (SPG63; MIM #615686) or pontocerebellar hypoplasia (PCH9; MIM #615809)^10–19^. Functional studies of *AMPD2* patient-derived cell lines and a murine model have suggested that shutdown of protein translation is the terminal pathogenic mechanism in AMPD2 deficiency, in line with other PCH-related genes^20,21^.

In this study, we report that bi-allelic, loss-of-function variants in *AMPD2* can cause Leigh syndrome at any life stage, without neurodevelopmental anomalies. Our findings expand the spectrum of AMPD2 deficiency to include severe mitochondrial dysfunction, likely resulting from disrupted cytosolic purine metabolism.

## Methods

### Participants

We recruited two adult patients (subjects A-II.4 and B-II.1 in Fig. 1A), clustered together via the White Matter (WM) Rounds Network (whitematterrounds.com)^22^, based on the following inclusion criteria: a) presence of bilateral, symmetric involvement of the striatal nuclei and brainstem on MRI, b) progressive neurological decline, and c) no molecular diagnosis despite extensive NGS panel-based clinical testing. We later identified two siblings from a third family (subjects C-II.1 and C-II.2 in Fig. 1A) through GeneMatcher^23^. Previously acquired clinical and radiological data were compiled and reviewed to orient the whole genome sequencing (WGS) analysis, as detailed below.

**Figure 1.** Segregation studies and neuroradiological features in three unrelated families with *AMPD2*-related Leigh syndrome.

### DNA extraction and WGS

Subject genomic DNA (gDNA) was isolated from whole blood using the QIAamp mini kit (Qiagen; 51104), diluted in nuclease-free water, and expedited for WGS to the Centre d’Expertise et de Services Genome Quebec (Montreal, QC). Paired-end, short-read WGS was performed on the NovaSeq6000 system (Illumina), reaching an average depth of coverage equal to 30X. Raw FASTQ read data generated from WGS were aligned to GRCh37 using the Burrows-Wheeler Aligner (BWA)^24^. Variant classes were then subjected to respective pipelines. Single-nucleotide variants (SNVs) and indels were called using the genome analysis toolkit (GATK) and annotated with ANNOVAR (with added in-house scripts)^25,26^. CNVkit was used to detect copy number variants (CNVs)^27^, ExpansionHunter Denovo was used to detect repeat expansions^28^, MToolBox was used to detect mtDNA alterations^29^, and the GATK Structural Variant (GATK-SV) pipeline was used for all other structural variants^25^. As an orthogonal approach, raw SNV and CNV .vcf files were supplementarily annotated and analyzed within the Franklin genome analysis platform (Qiagen; https://franklin.genoox.com). We filtered variants based on rarity and category of the harbouring gene, in the following order of priority: 1) associated with mitochondrial disease^30^, 2) encoding a mitochondria-localized protein^31^, 3) shared between subjects A-II.4 and B-II.1, 4) associated with leukoencephalopathy^30^, 5) associated with neurogenetic disease^30^, 6) containing two or more rare, predicted deleterious (Franklin aggregate score ≥0.7) variants. Sanger sequencing for variant validation and segregation studies was done on the 3730xl DNA Analyzer from Applied Biosystems (Genome Quebec). Primers were designed using the Primer-BLAST web tool to ensure specificity, and amplicons encapsulating variants of interest were generated through touchdown PCR with standard reagents^24^. Chromatograms and genotype-phenotype correlations were interpreted in SnapGene (Dotmatics).

### Cell culture

Mutant primary fibroblasts/myoblasts were cultivated from dermal/muscular biopsies following informed consent and immortalized, as previously described^32–34^. Stable isogenic controls (AMPD2 overexpression) were engineered via retroviral transduction (DNASU; plasmid HsCD00505768), as previously described^32,33,35^. Myoblasts at 90% confluence were differentiated into myotubes upon 72hr exposure to fusion medium, as previously described^36^. For adenosine challenge experiments, cells were first seeded to 90% confluence in full medium. Then, cells/myotubes were washed twice with phosphate-buffered saline (PBS) prior to application of serum-free/fusion medium with or without 500μM adenosine (Thermo) for 72hr.

Medium was replaced daily – adenosine was prepared fresh just before replacement. All cell culture incubation was at 37°C in a 5% CO_2_ atmosphere, and all cultures were ensured as devoid of contamination, including mycoplasma.

- Immortalized skin fibroblast full medium: DMEM X-Cell (Wisent; 319-027-CL) + 10% Fetal bovine serum (FBS) (Wisent; 098-150) + penicillin-streptomycin (Wisent; 450-201-EL).
- Immortalized myoblast full medium: SKMax (Wisent; 301-060-CL) + 10% FBS + SKMax supplement (Wisent; 301-061-XL) + penicillin-streptomycin.
- Fusion medium: DMEM X-Cell + 2% heat-inactivated horse serum (Wisent; 065-210) + penicillin-streptomycin.

### Immunoblotting

For SDS-PAGE, cells were harvested in PBS after trypsinization, pelleted (centrifuged at 1000 x g for 5min), then lysed on ice for 20min in 1.5% n-dodecyl β-D-maltoside (DDM; in PBS) supplemented with protease inhibitors (Roche; 11836170001). Samples were then centrifuged at 4°C (20 000 x g) for 20min, after which 30μg of protein from the supernatant was quantified by Bradford assay (Bio-Rad; for cell isolates) or bicinchoninic acid (BCA) assay (Thermo; for tissue extracts), prepared in Laemmli buffer, denatured (50°C for 10min), and loaded onto 8-12% polyacrylamide gels. BN-PAGE was performed on 6–15% gradient polyacrylamide gels, using 30μg of isolated mitochondria obtained from patient skeletal muscle or mitoplasts from myoblast/fibroblast lines, as previously described^33,37^. Resolved proteins were transferred to nitrocellulose membranes and probed indirectly: overnight primary antibody incubation at 4°C, three-time PBS wash (30min), 2hr incubation with horseradish peroxidase-conjugated secondary antibodies (Jackson ImmunoResearch Labs), and a final, three-time PBS wash (30min). Membranes, post-application of chemiluminescence reagent (Cell Signaling Technology; 7003S), were imaged using the Chemostar system (Intas).

### Slide preparation for immunofluorescence

Cells were seeded in 24-well plates containing coverslips and fixed in 4% paraformaldehyde (in PBS) immediately after experimental termination for 20min. Coverslips were washed three times with PBS and then treated with 0.1% Triton X-100 (in PBS) for 15min to allow permeabilization. After another three PBS washes, coverslips were blocked in 5% bovine serum albumin (BSA; in PBS) for 30min and subsequently incubated with a primary antibody cocktail prepared in 5% BSA for 1.5hr. Coverslips were washed again three times with PBS and incubated with an appropriately configured, Alexa fluorophore-conjugated (Invitrogen) secondary antibody cocktail for 30min. After a final three PBS washes, coverslips were placed securely on slides using mounting medium (Dako), incubated at 37°C for 10min, and chilled at -20°C for at least 20min prior to visualization.

### Confocal microscopy

The imaging apparatus used was an Olympus IX81 inverted microscope with accompanying Andor/Yokogawa spinning disk system (CSU-X) and sCMOS camera. Prepared coverslips were visualized at 60X magnification (NA1.4 lens). 150 Z-stack images (11 steps, 0.2μm/step) of non-overlapping cells were acquired per experimental condition to enable quantitative analysis of mitochondrial network morphology using the ImageJ plugin MitochondriaAnalyzer (https://github.com/AhsenChaudhry/Mitochondria-Analyzer)^38^. Briefly, the channels corresponding to the mitochondrial probe (anti-PRDX3) were isolated and converted to 8-bit TIFF format using the built-in ImageJ batch-convert function (Process>Batch>Convert). Compatible files were then batch-analyzed after the scale was set using the 3D batch command option in MitochondriaAnalyzer (Plugins>MitochondriaAnalyzer>3D>3D Batch Commands). The default values for thresholding parameters were applied consistently to all images analyzed. Quantification of IMPDH2 oligomers was performed by visual inspection. Three biological replicates were represented in the quantification datasets per condition (mitochondrial morphology = 50 field images/replicate; IMPDH2 oligomers = 50 visually delineated single cells/replicate).

### Proteomics

50μg of isolated mitochondria from skeletal muscle (derived from healthy control and subject B-II.1) were solubilized in 50μL (1μg/μL) 2% SDS (in PBS) for 20min and pelleted (20000 x g for 20min). The entire supernatant was then expedited for liquid chromatography mass spectrometry (LC-MS), conducted at the Institut de recherches cliniques de Montréal LC-MS platform. Mascot scores generated for each unique peptide detected were averaged across two technical replicates, and the fold changes (patient versus control) were computed. Mutually exclusive peptides detected between patient and control were assigned a mascot score of 0.1 in the opposing dataset (e.g., a peptide detected in the patient but not in the control was entered manually into the control dataset and assigned a mascot value equal to 0.1). Gene ontology (GO) term enrichment analysis was performed on proteins with fold changes >1.25 or <0.75 using Enrichr^39^.

### Primary antibodies

AMPD2 (Santa Cruz; sc-100504), IMPDH2 (Abcam; ab129165), HSP60 (Santa Cruz; sc-376261), PRDX3 (in-house), SDHA (Abcam; ab110252), ATP5F1A (Abcam; ab14748), NDUFA9 (Abcam; ab14713), UQCRC1 (Abcam; ab14745), alpha-Tubulin (ProteinTech; 66031-1-Ig), beta-Actin (GenScript; A00702), COX4 (in-house).

### Statistical analyses

Data were entered and analyzed using Prism (GraphPad). For mitochondrial morphology and IMPDH2 metrics, statistical significance was evaluated through Kruskal-Wallis test followed by Dunn’s multiple comparisons tests if the data failed normality/homogeneity of variance tests and/or if outliers were present. Otherwise, one-way ANOVA followed by Tukey’s post-hoc analysis was performed. An adjusted p-value of less than 0.05 was considered statistically significant.

### Ethical approval

The research studies on cell lines were approved by the institutional review board of The Neuro (Montreal Neurological Institute-Hospital), McGill University. Informed consent was obtained from all subjects or parents/legal guardian.

## Results

### Bi-allelic variants in AMPD2 cause Leigh syndrome with variable onset and severity

Extensive clinical genetic testing including NGS-based targeted panels for genetic leukoencephalopathies, mitochondrial disorders, and hereditary neuropathies were inconclusive for both unrelated adult male subjects. Therefore, we performed research-level WGS in search of candidate disease-causing variants. The WGS analysis returned no potential molecular diagnosis across nuclear- and mtDNA-encoded mitochondrial disease genes. Yet, it revealed that each subject carried two extremely low allele frequency variants in the *AMPD2* gene in the heterozygous state. Each *AMPD2* variant was either annotated as loss-of-function or missense with unanimously deleterious predictions across *in silico* tools (Fig. 2A). Sanger sequencing demonstrated that both sets of variants were in trans and co-segregated with the disorder in each family, consistent with autosomal recessive inheritance (Fig. 1A). For both these cases, one variant was located in the second coding exon (not present in all AMPD2 isoforms), while the other was located in an exon corresponding to the deaminase domain of the enzyme (Fig. 2B).

**Figure 2.**
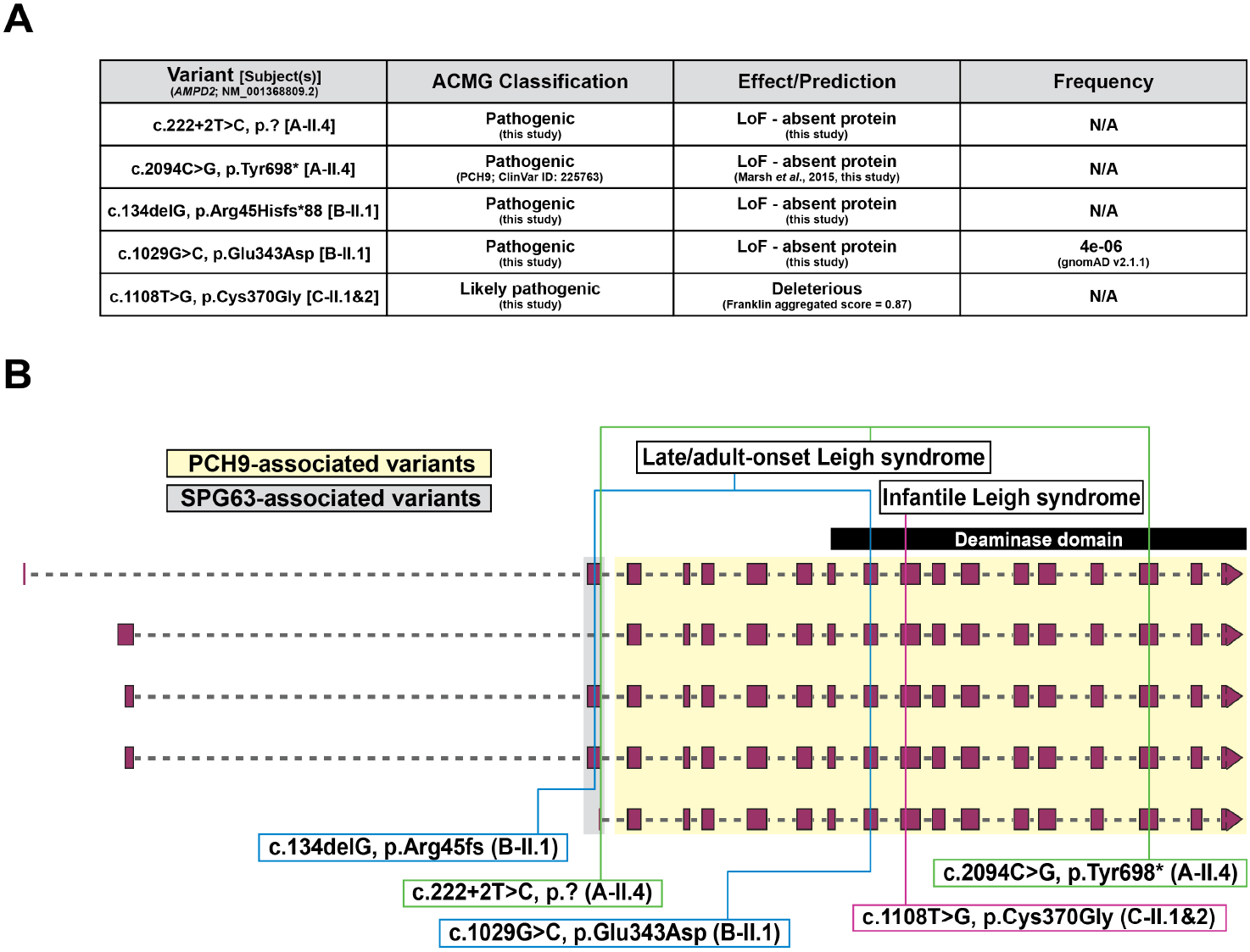
*AMPD2* variant classifications and genotype-phenotype relationships. (A) ACMG classifications, predictions, and allele frequencies of bi-allelic *AMPD2* variants detected. (B) Genotype-phenotype correlations found in this study, labeled by subject. ACMG, American College of Medical Genetics; LoF, loss-of-function; N/A, not available; PCH9, pontocerebellar hypoplasia type 9; SPG63, spastic paraplegia type 63.

After establishing *AMPD2* as the likely causal gene in our first two unrelated adult Leigh syndrome subjects, we identified a third *AMPD2*-Leigh syndrome (*AMPD2*-LS) family through GeneMatcher^23^. Two affected males, one child and one adult – both with onset at birth, were found to be homozygous for a predicted deleterious, missense, deaminase domain variant (Fig. 2). The variant was detected by exome sequencing after substantial clinical genetic workup and co-segregated with the disorder in the family. An unaffected sibling, a year younger than the affected child, was established as a carrier for the variant (Fig. 1A).

### AMPD2 loss-of-function coincides with mitochondrial dysfunction

To determine the effect of the variants detected on protein function, we generated skin fibroblast and skeletal myoblast cultures from patient-derived dermal/muscular biopsies. The mutant fibroblast line from subject A-II.4, the mutant fibroblast and myoblast lines from subject B-II.1, and skeletal muscle from subject B-II.1 all showed total loss of AMPD2 protein and native AMPD2 complex (Fig. 3A), validating a loss-of-function impact and ascribing full pathogenic status to their *AMPD2* variants.

**Figure 3.**
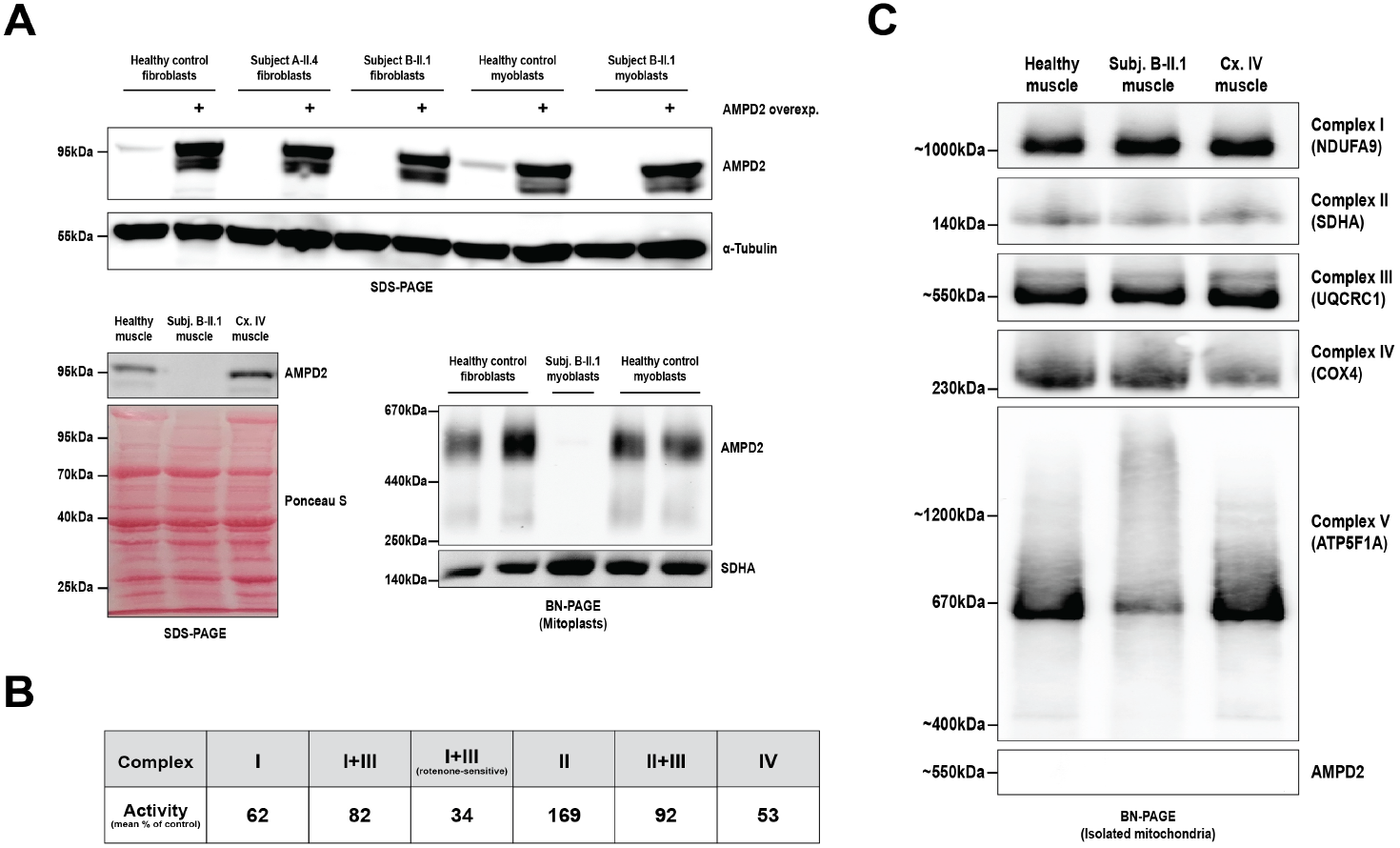
Biochemical diagnosis of concomitant AMPD2 deficiency and Leigh syndrome. (A) Immunoblot analyses validating loss-of-function effect for *AMPD2* variants detected. Top: SDS-PAGE of whole cell lysates from control and patient-derived cell lines with or without retroviral transduction of AMPD2 overexpression construct. Bottom-left: SDS-PAGE of healthy control-, subject B-II.1-, and complex IV (Cx. IV)-related Leigh syndrome patient (positive control)-derived skeletal muscle cytoplasmic fractions. Bottom-right: BN-PAGE indicating loss of ∼550kDa native AMPD2 complex in a patient-derived myoblast line. (B) Respiratory chain enzymatic activity profile of subject B-II.1 skeletal muscle mitochondria, after normalization to citrate synthase activity, reported as mean % of control values (two trials). All values were >30%, unsatisfactory for the diagnosis of a respiratory chain disorder according to the modified Walker criteria^40^. (C) BN-PAGE of skeletal muscle mitochondria providing diagnostic confirmation of complex V-related Leigh syndrome in our *AMPD2* subject and absence of AMPD2 protein in mitochondrial fractions.

To provide evidence of mitochondrial dysfunction, we first expedited a skeletal muscle specimen (subject B-II.1) for mitochondrial respiratory chain enzymatic activity analyses (Baylor Genetics). This test showed diminished complex I and complex IV activities (normalized to citrate synthase) that were interpreted as non-diagnostic (>30% residual activity) for a respiratory chain disorder according to the modified Walker criteria (Fig. 3B)^40^. We then performed blue native polyacrylamide gel electrophoresis (BN-PAGE) on mitochondria isolated from skeletal muscle to screen for respiratory complex assembly defects. The BN-PAGE unveiled a severe abrogation of complex V (ATP synthase) formation with accumulation of high molecular weight oligomers, while complexes I through IV appeared normal (Fig. 3C). These results demonstrate, for the first time, that loss of AMPD2 profoundly disrupts OXPHOS through complex V assembly, resulting in LS.

Because of the muscular pathology, the phenotypic spectrum now associated with *AMPD2*, and the specific role of AMPD2 in purine metabolism, we hypothesized that a unique mechanism underlies mitochondrial dysfunction in AMPD2 deficiency. To gain insight, we performed proteomic analyses of healthy control- and *AMPD2* subject (B-II.1)-derived, mitochondria-enriched skeletal muscle samples. Firstly, we validated that no AMPD2 peptides were detected in the proteomics data (eTable 2) and that the protein could only be detected in control skeletal muscle cytosolic but not mitochondrial fractions (Fig. 3A, C). This eliminated the possibility of mitochondrial localization and direct contribution to OXPHOS. Targeted analysis of complex V subunit and assembly factor abundances pinpointed the loss of subunit s (also known as factor B) and assembly factor FMC1 in our *AMPD2* subject (Fig. 4A). Gene ontology (GO) term enrichment analysis principally showed upregulation of cytosolic translation and glycolysis and downregulation of many critical mitochondrial processes in our *AMPD2* subject versus healthy control (Fig. 4B). In brief, this profile highlighted a multifaceted mitochondrial impairment associated with significant metabolic decompensation that aligned with LS.

**Figure 4.**
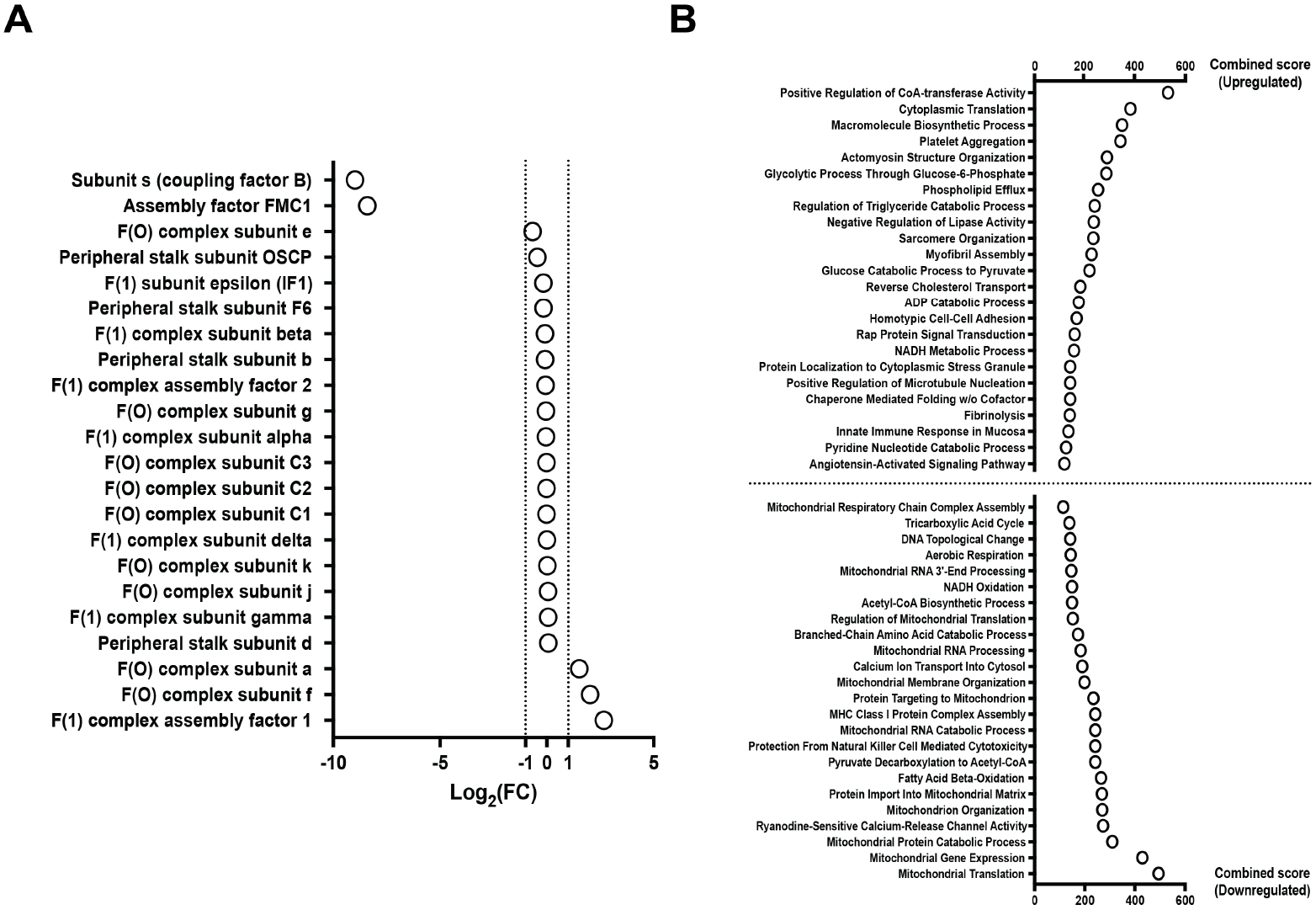
Proteomic analysis of AMPD2-null skeletal muscle mitochondria. (A) Protein score fold changes (FC; subject B-II.1 vs. healthy control) of respiratory complex V subunits detected by LC-MS. (B) Gene ontology (GO)-term enrichment analysis of differentially regulated biological processes from the proteomics data, generated using Enrichr. The combined score reflects both the degree of statistical significance and overrepresentation of a biological process in the top 24 significantly upregulated (top) or top 24 downregulated (bottom) gene lists^39^. All processes listed had a corresponding adjusted p<0.05.

### Adenosine overload promotes mitochondrial hyperfusion in AMPD2-knockout cells

Adenosine-loaded, serum-free medium, rather than full cell culture medium, was previously shown to induce translation inhibition and cell death in AMPD2-knockout cells derived from PCH9 patients (Fig. 5A)^17^. Hence, we tested whether mitochondrial dysfunction was present under these conditions. Because cellular GTP is pathologically low in this model, the main proteins influencing mitochondrial dynamics are GTPases, and dysregulation of mitochondrial fission/fusion balance is a recognized pathomechanism in PCH, we first reasoned that the mitochondrial network would be affected^33^. We thus utilized immunofluorescence confocal microscopy to quantify the state of the mitochondrial network in challenged cells. Compared to AMPD2-expressing lines, mitochondria in the mutant lines demonstrated reduced sphericity, increased branch numbers, junctions, and end points, and increased mean and total branch lengths, indicating network hyperfusion (Fig. 5B, eFigure 1).

**Figure 5.**
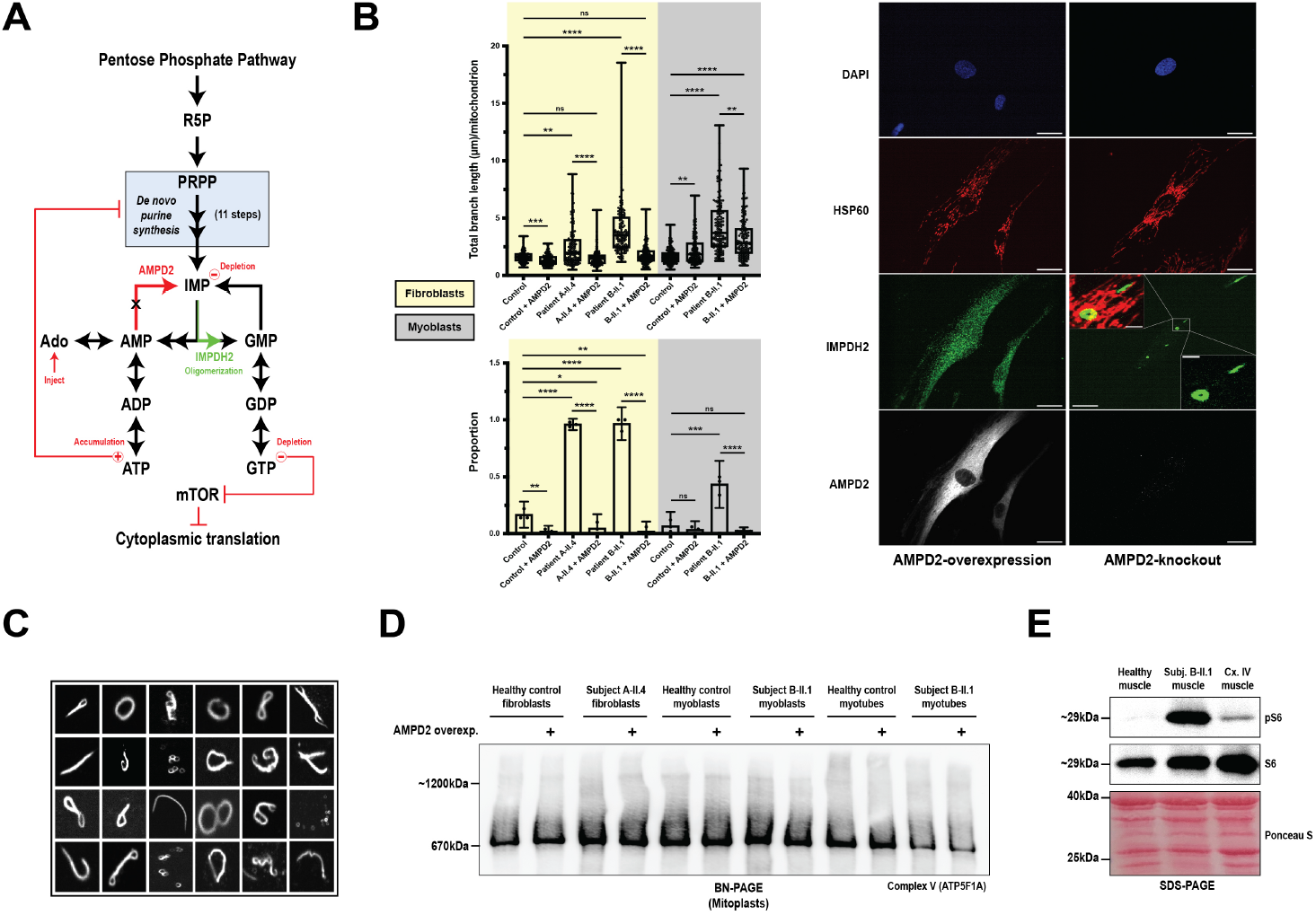
Assessment of mitochondrial dysfunction in challenged AMPD2-null cells. (A) Schematic of the proposed pathogenic mechanism in AMPD2 deficiency and rationale for adenosine (Ado) challenge^17,21^. (B) Top left: quantification of total mitochondrial object length across cell lines treated with adenosine (n = 150 cells per line; three biological replicates of 50 cells pooled). Whiskers denote minima and maxima. Bottom left: proportion of cells containing at least one IMPDH2 oligomer (three biological replicates of 50 cells). Bars indicate mean +/- 95% confidence intervals. Right side: representative confocal micrographs of challenged AMPD2-overexpressing and AMPD2-knockout lines. HSP60 was used as the mitochondrial probe. Scale bars at the bottom of each image correspond to a distance of 20μm. Scale bars in close-up panels indicate a distance of 3μm. (C) Collage of high-order IMPDH2 oligomeric forms visualized in challenged mutant cells. (D) BN-PAGE of mitoplasts derived from challenged cells and myotubes indicating no defect in complex V assembly. (E) SDS-PAGE of skeletal muscle cytosolic fractions showing heavy mTOR activation in AMPD2-null muscle compared to healthy and complex IV (Cx. IV)-deficient muscle, proxied by phosphorylated ribosomal protein S6 (pS6) levels. *p<0.05; **p<0.01; ***p<0.001; ****p<0.0001; ns, not significant (p>0.05).

### Mutant cells show dramatic IMPDH2 oligomerization during adenosine challenge

Oligomerization of the enzyme IMPDH2, known to occur when cells exhibit an elevated ATP/IMP:GTP ratio, was previously associated with neuroprotection in *Ampd2*/*Ampd3* double knockout (dKO) mouse brain and in patient-derived neural progenitor cells (NPCs) under baseline culture conditions^17,21^. Therefore, we stained our challenged myoblasts and fibroblasts for IMPDH2 oligomers to replicate and confirm, by proxy, that only mutant lines were suffering from major purine nucleotide imbalance. Indeed, only cell lines lacking AMPD2 demonstrated robust micron-sized IMPDH2 oligomerization in response to adenosine treatment, most commonly in the form of rods or rings that typically did not colocalize with mitochondria (Fig. 5B, eFigure 2). The oligomers, however, appeared in a variety of high-order shapes and sizes in addition to common rod- or ring-like configurations (Fig. 5C, eFigure 2). Oligomerization in mutant lines was not associated with an increase in IMPDH2 protein levels, and IMPDH2 levels were also not increased in skeletal muscle from subject B-II.1 (eFigure 3).

### Stressed mutant cells and myotubes do not recapitulate skeletal muscle pathology

Since AMPD2-null cell lines exhibited striking phenotypes upon adenosine challenge, we tested the broader validity of this experimental condition for the study of *AMPD2*-LS. BN-PAGE revealed that adenosine challenge was not sufficient to induce the complex V assembly defect previously detected in patient skeletal muscle (Fig. 5D). To potentially recreate the complex V defect, we differentiated myoblasts into myotubes and subjected these to adenosine challenge. This also failed to reproduce the defect (Fig. 5D). Following our GO-term analysis, which exposed cytosolic translation as the upregulated process with highest number of contributing genes, we probed for mTOR activation by measuring levels of total and phosphorylated ribosomal protein S6 (pS6). Although pS6 and total S6 levels remained unchanged across challenged cell lines (eFigure 3), pS6 levels were drastically increased in patient skeletal muscle compared to controls (Fig. 5E).

## Discussion

In this study, we show that bi-allelic, pathogenic *AMPD2* variants can cause variable-onset LS. Thus, we expand the spectrum of AMPD2 deficiency, demonstrating that symptoms can appear at any age without neurodevelopmental anomalies. The evidence we provided supports a mitochondrial pathogenesis of AMPD2 deficiency, shedding new light on the interconnectedness between cytosolic purine metabolism and OXPHOS.

### International collaboration and WGS

This work exemplifies how international case-based discussions and data sharing can reduce diagnostic odysseys of undiagnosed patients with ultra-rare genetic diseases. All subjects here included – from different ethnic backgrounds and geographic origins – benefitted considerably from the WM Rounds Network and GeneMatcher^22,23^. Specifically, the WM Rounds Network enabled access to a validated research-level WGS pipeline that eventually led to molecular diagnosis. This vital step permitted downstream functional study of the disease, ameliorating our understanding of fundamental metabolic processes.

### Prior evidence for mitochondrial disease

Indicators of mitochondrial dysfunction have been sporadically reported in *AMPD2*-PCH9 human patients: small, bilaterally hyperintense deep grey matter nuclei in one family^19^, complex I deficiency by enzymology – interpreted as a potential false positive result – in a four-year-old female^18^, and elevated lactate on brain magnetic resonance spectroscopy in another male infant^14^. Moreover, dogs with a homozygous deaminase domain frameshift mutation in the canine ortholog of *AMPD2* showed extrapyramidal movement disorders with bilateral white matter and caudate nuclei signal abnormalities^41^. Taken with the results of our study, these findings constitute a body of evidence now pointing to an evolutionary connection between AMPD2 activity and OXPHOS.

### AMPD2 and mtDNA

Currently, 134 nuclear- and 14 mtDNA-encoded genes are causally associated to LS^42^. Over 98% of these genes encode products that localize to mitochondria and directly participate either in the formation of electron transport chain complexes, mtDNA replication/maintenance, membrane dynamics, cofactor biosynthesis, or B vitamin/organic acid metabolism^3,4,42^. However, two of these genes encode cytosolic enzymes implicated in nucleotide metabolism, like *AMPD2*: *TYMP* and *RRM2B*. When mutated, however, both genes result in nucleotide imbalance and mtDNA alteration/depletion syndromes with hallmark findings of combined OXPHOS deficiency and ragged red fibers^43–45^. While we did not evaluate mtDNA content, the BN-PAGE respiratory complex assembly profile, respiratory enzymatic activity analyses, and lack of ragged red fibers in our subjects’ skeletal muscle together suggest that mitochondrial dysfunction in AMPD2 deficiency is not resulting from mtDNA alteration/depletion but rather a unique pathological mechanism acting specifically on complex V.

### Comparison to known models of AMPD2 deficiency

Whole brains of dKO mice as well as challenged AMPD2-knockout neuronal and non-neuronal cell lines were all shown to exhibit simultaneous ATP accumulation and GTP deficiency^17^. Therefore, by virtue of identifying *AMPD2*-LS patients, this work represents a fascinating, paradoxical instance whereby AMPD2 deficiency, a condition in which cellular ATP concentrations are pathologically elevated, nevertheless causes LS, a disorder traditionally ascribed to failure of ATP production.

Secondly, whereas the dKO mice displayed robust upregulation and oligomerization of IMPDH2 in affected brain regions compared to controls, challenged AMPD2-knockout myoblasts and fibroblasts demonstrated abundant IMPDH2 oligomerization without upregulation^21^. IMPDH2 upregulation was neither seen in skeletal muscle from subject B-II.1 (oligomerization status unknown). These data indicate that IMPDH2 regulation is likely disparate across tissues, perhaps conferring differential susceptibility to nucleotide imbalance.

Lastly, whereas the dKO mouse, challenged AMPD2-knockout NPCs, and challenged *amd1* mutant yeast demonstrated shutdown of protein translation (greatly reduced pS6 in dKO and less ^35^S-methionine incorporation in challenged mutant NPCs/yeast), we detected 1) no changes in pS6 levels in challenged myoblasts and fibroblasts and 2) highly elevated pS6 levels in patient skeletal muscle versus controls^17,21^. The translation phenotype in human AMPD2-null skeletal muscle is therefore the only phenotype that remains consistent with many well-characterized cellular and animal models of LS^46–48^. Since the complex V assembly defect and elevated pS6 levels from patient skeletal muscle could not be found in challenged mutant fibroblasts and myoblasts, which showed mitochondrial hyperfusion and blatant IMPDH2 oligomerization, it remains plausible that IMPDH2 oligomerization protects against OXPHOS failure and eventual cell death. Deeper study of AMPD2-knockout models is necessary to find the metabolic conditions that trigger simultaneous complex V deficiency and mTOR activation.

### No paralog compensation

Intriguingly, the paralogs *AMPD1, AMPD2*, and *AMPD3* are all expressed in human skeletal muscle, and the corresponding encoded enzymes are each believed to exhibit AMP deaminase activity^20^. AMPD1 exhibits tissue specialization for skeletal muscle and is the highest expressed paralog in this tissue, followed by AMPD2 and AMPD3 (lowest)^20^. However, we show that loss of *AMPD2* alone is sufficient to induce a severe complex V assembly defect and metabolic derailment in this tissue. These findings indicate that *AMPD* paralogs likely either have divergent functions that go beyond deaminase activity or substrate flux/efficiencies that do not positively correlate with protein levels.

### Likely disruption of mitochondrial cristae

Our proteomic analysis revealed that two matrix-localized, soluble components of complex V were missing from *AMPD2* patient skeletal muscle: coupling factor b (ATP synthase subunit s) and protein FMC1. Factor b is posited to help maximize complex V efficiency through blocking a proton leak within ribbons of ATP synthase dimers, which are found at highly curved apices of the mitochondrial cristae^49–56^. FMC1, conversely, participates redundantly in ATP synthase assembly at baseline conditions, only becoming essential under heat stress conditions^57^. Since AMPD2 loss results in purine nucleotide imbalance where ATP levels increase while GTP levels decrease, and complex V assembly is affected in isolation, we hypothesize that the scarcity of GTP hampers proper OPA1-mediated cristae junction formation and maintenance^58^. Dysmorphic cristae in this context may promote aberrant ATP synthase oligomerization and dissociation of soluble ATP synthase components, explaining the BN-PAGE profile and decreased levels of factor b and FMC1. Although we did not visualize cristae in this study, the muscle proteomics data did not indicate an overt reduction of any particular cristae-localized protein.

## Conclusions

The novel identification of *AMPD2*-LS as a clinical entity broadens the spectrum of *AMDP2*-related conditions and points to an evolutionary interdependence between cytosolic purine metabolism, complex V formation, OXPHOS, and the structural integrity of mitochondrial cristae. Our findings raise the question of whether purine levels could be modulated therapeutically for the treatment of PCH, LS, and other neurological disorders in which mitochondria play a central pathological role. To this end, further study of AMPD2 deficiency will aid to elucidate how inhibitors of mTOR and cyclic GMP degradation (rapamycin and sildenafil respectively) act to extend lifespan and/or improve outcomes in LS models and human subjects^46–48,59^. Finally, the diagnostic resolution of this case series through WGS illustrates that predicted deleterious/loss-of-function variants in known Mendelian genes presently associated with uniformly devastating, infantile phenotypes should not be deprioritized when found in mildly affected adults.

## Data Availability

All data in the present study are available upon reasonable request to the authors.

---

**eTable 1. Summary of clinical, radiological, and molecular findings in *AMPD2*-LS subjects**

**eFigure 1.**
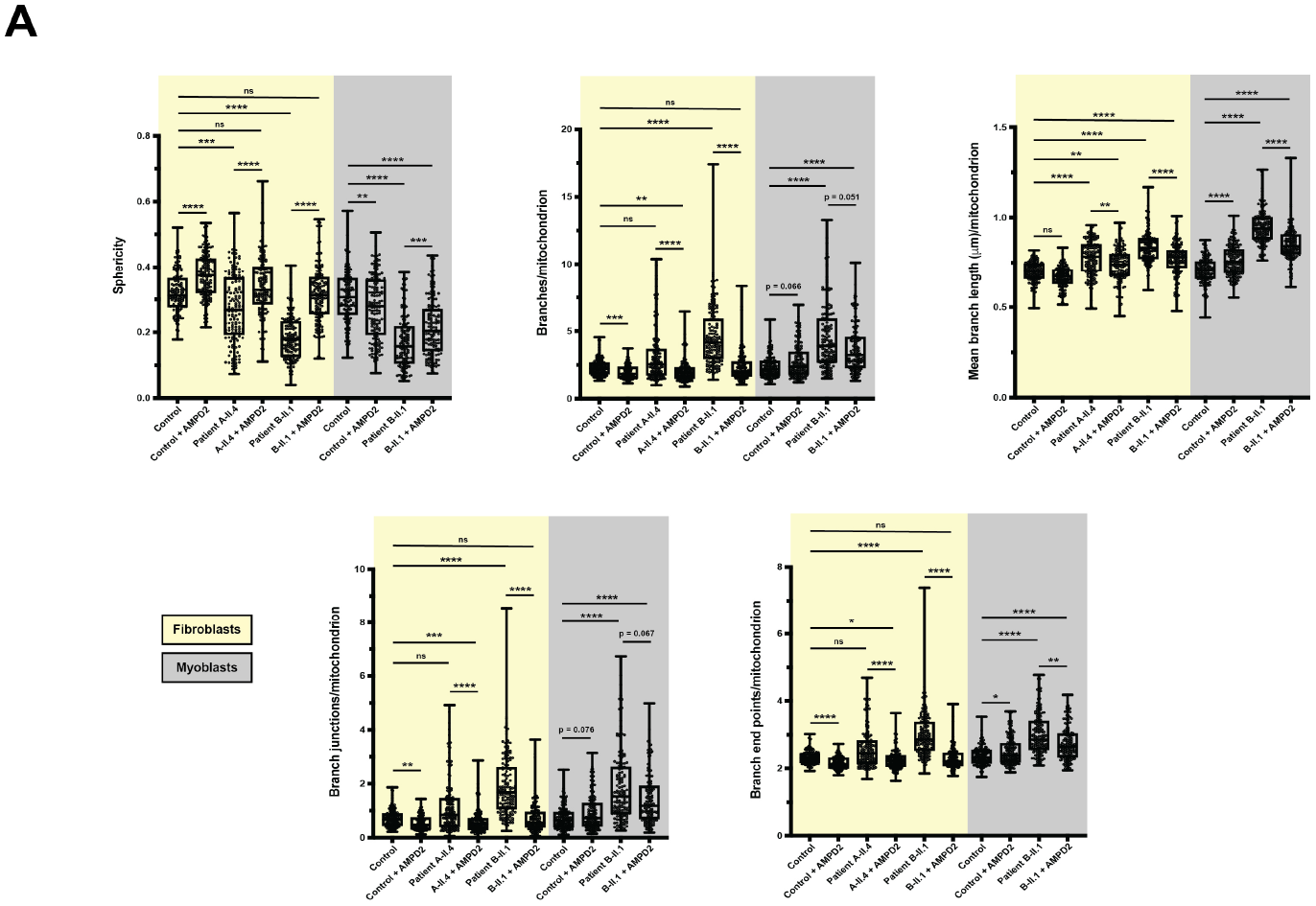
Quantitative analysis of mitochondrial morphology parameters (related to Fig. 5) (A) Box and whisker plots of mitochondrial morphology parameters across cell lines treated with adenosine (n = 150 cells per line; three biological replicates of 50 cells pooled), which together indicate a hyperfused state of the mitochondrial network in mutant lines. Whiskers denote minima and maxima. *p<0.05; **p<0.01; ***p<0.001; ****p<0.0001; ns, not significant (p>0.05).

**eFigure 2.**
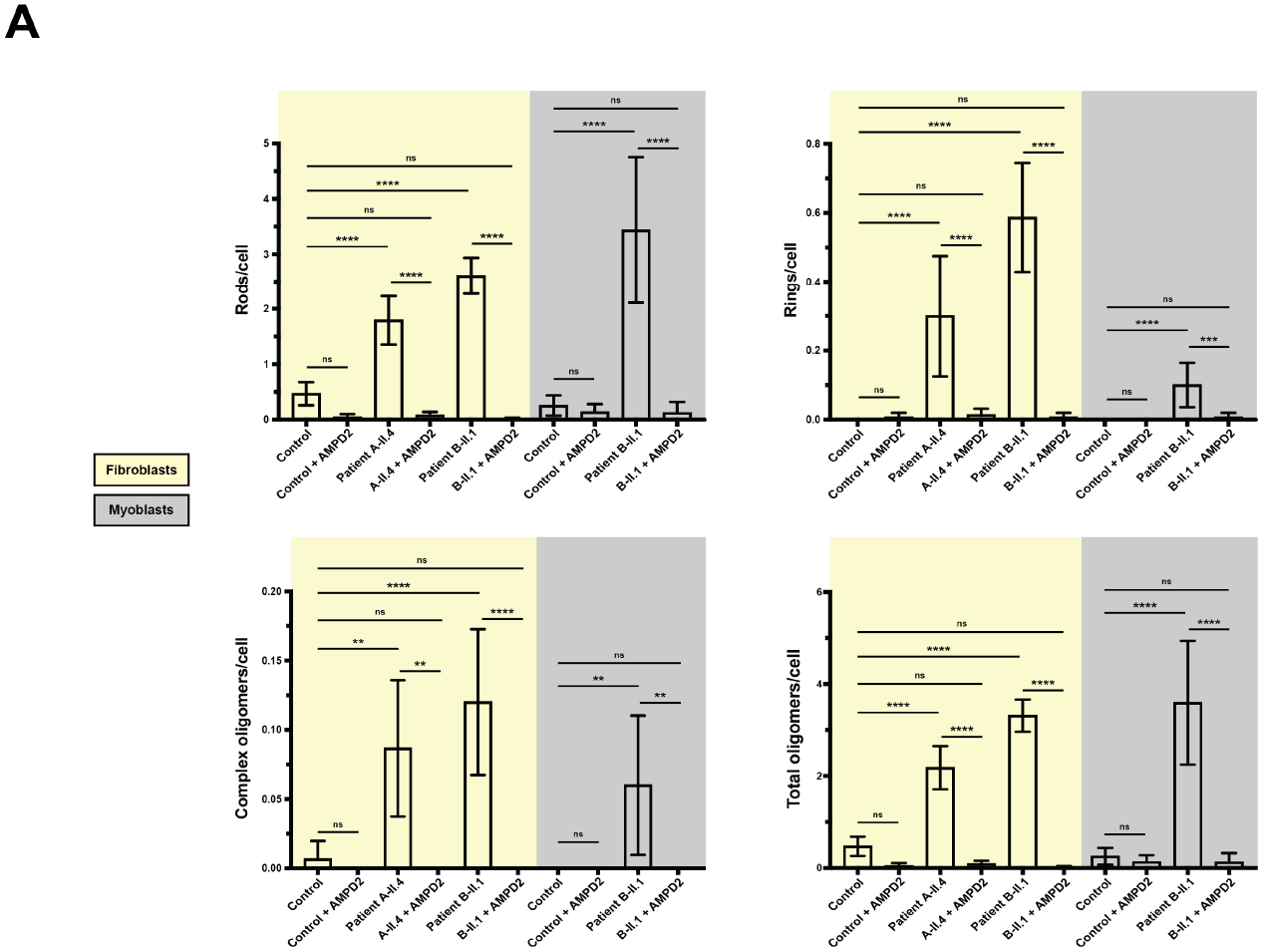
Quantification of IMPDH2 oligomeric forms (related to Fig. 5) (A) Bar plots displaying the relative amounts of different IMPDH2 oligomeric forms across cell lines treated with adenosine (n = 150 cells per line; three biological replicates of 50 cells pooled). IMPDH2 oligomers were almost exclusively found in mutant lines. Bars indicate mean +/- 95% confidence intervals. *p<0.05; **p<0.01; ***p<0.001; ****p<0.0001; ns, not significant (p>0.05).

**eFigure 3.**
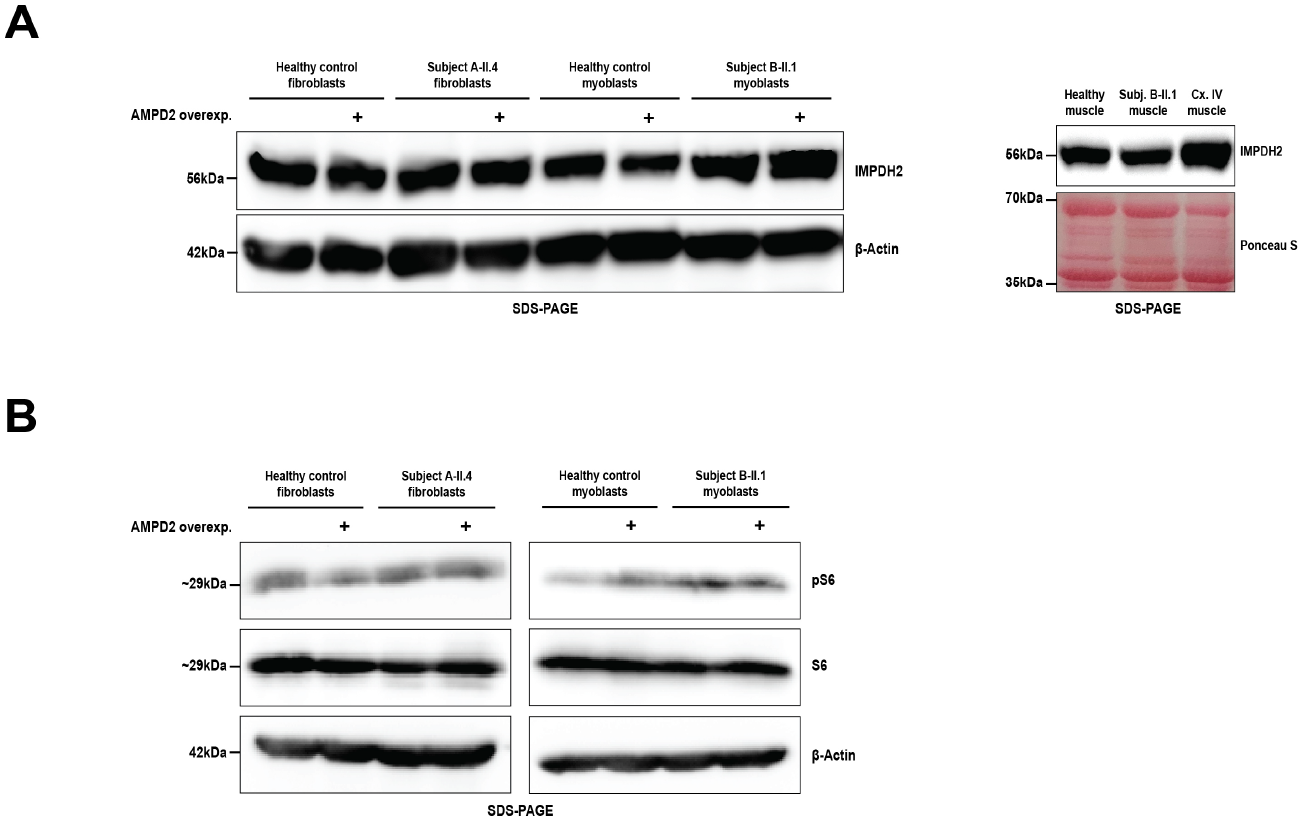
Immunoblot analysis of IMPDH2 and phosphorylated S6 levels. (A) Left: immunoblot of cell lines treated with adenosine indicating no change in IMPDH2 levels. Right: immunoblot of skeletal muscle cytoplasmic fractions indicating no change in IMPDH2 levels across healthy control, AMPD2-null (subject B-II.1), and complex IV (Cx. IV)-deficient muscle. (B) Immunoblot of cell lines treated with adenosine indicating no change in pS6/S6 levels.

